# Longitudinal immune profiling reveals distinct features of COVID-19 pathogenesis

**DOI:** 10.1101/2020.06.13.20127605

**Authors:** Elizabeth R. Mann, Madhvi Menon, Sean Blandin Knight, Joanne E. Konkel, Christopher Jagger, Tovah N. Shaw, Siddharth Krishnan, Magnus Rattray, Andrew Ustianowski, Nawar Diar Bakerly, Paul Dark, Graham Lord, Angela Simpson, Timothy Felton, Ling-Pei Ho, Respiratory TRC, Marc Feldmann, CIRCO, John R. Grainger, Tracy Hussell

**Author notes:** Joint corresponding authors, Correspondence: Professor Tracy Hussell or Dr John Grainger. contributed equally (Elizabeth R. Mann, Madhvi Menon, Sean Blandin Knight, Joanne E. Konkel, John R. Grainger, Tracy Hussell). CIRCO investigators: Rohan Ahmed, Halima Ali Shuwa, Miriam Avery, Katharine Birchall, Oliver Brand, Evelyn Charsley, Alistair Chenery, Christine Chew, Richard Clark, Emma Connolly, Karen Connolly, Simon Dawson, Laura Durrans, Hannah Durrington, Jasmine Egan, Claire Fox, Helen Francis, Miriam Franklin, Susannah Glasgow, Nicola Godfrey, Kathryn J. Gray, Seamus Grundy, Jacinta Guerin, Pamela Hackney, Mudassar Iqbal, Chantelle Hayes, Emma Hardy, Jade Harris, Anu John, Bethany Jolly, Verena Kästele, Saba Khan, Gabriella Lindergard, Sylvia Lui, Lesley Lowe, Alex G Mathioudakis, Flora A. McClure, Joanne Mitchell, Clare Moizer, Katrina Moore, David Morgan, Stuart Moss, Syed Murtuza Baker, Rob Oliver, Grace Padden, Christina Parkinson, Laurence Pearmain, Mike Phuychareon, Ananya Saha, Barbora Salcman, Nicholas A. Scott, Seema Sharma, Jane Shaw, Joanne Shaw, Elizabeth Shepley, Lara Smith, Simon Stephan, Ruth Stephens, Gael Tavernier, Rhys Tudge, Louis Wareing, Roanna Warren, Thomas Williams, Lisa Willmore, Mehwish Younas. NIHR Respiratory TRC: Alex Horsley, Manchester BRC Tim Harrison, Nottingham BRC Joanna Porter, UCL BRC Ratko Djukanovic, Southampton BRC Stefan Marciniak, Cambridge BRC Chris Brightling, Leicester BRC Ling-Pei Ho, Oxford BRC Lorcan McGarvey, Queen’s University Belfast Jane Davies, Imperial College BRC.

## Abstract

**Background:** The pathogenesis of COVID-19, caused by a novel strain of coronavirus (SARS-CoV-2), involves a complex host-virus interaction and is characterised by an exaggerated immune response, the specific components of which are poorly understood. Here we report the outcome of a longitudinal immune profiling study in hospitalised patients during the peak of the COVID-19 pandemic in the UK and show the relationship between immune responses and severity of the clinical presentation.

**Methods:** The Coronavirus Immune Response and Clinical Outcomes (CIRCO) study was conducted at four hospitals in Greater Manchester. Patients with SARS-CoV-2 infection, recruited as close to admission as possible, provided peripheral blood samples at enrolment and sequentially thereafter. Fresh samples were assessed for immune cells and proteins in whole blood and serum. Some samples were also stimulated for 3 hours with LPS and analysed for intracellular proteins. Results were stratified based on patient-level data including severity of symptoms and date of reported symptom onset.

**Findings:** Longitudinal analysis showed a very high neutrophil to T cell ratio and abnormal activation of monocytes in the blood, which displayed high levels of the cell cycle marker, Ki67 and low COX-2. These properties all reverted in patient with good outcome. Unexpectedly, multiple aspects of inflammation were diminished as patients progressed in severity and time, even in ITU patients not recovering.

**Interpretation:** This is the first detailed longitudinal analysis of COVID-19 patients of varying severity and outcome, revealing common features and aspects that track with severity. Patients destined for a severe outcome can be identified at admission when still displaying mild-moderate symptoms. We provide clues concerning pathogenesis that should influence clinical trials and therapeutics. Targeting pathways involved in neutrophil and monocyte release from the bone marrow should be tested in patients with COVID-19.

**Funding:** The Kennedy Trust for Rheumatology Research, The Wellcome Trust, The Royal Society, The BBSRC, National Institute for Health Research (NIHR) Biomedical Research Centres (BRC).

**Research in context:** *Evidence before this study:* Analysis of the literature before the study via pubmed and bioRxiv searches using the terms COVID-19, SARS-CoV2, immune and inflammation (with the last search performed on 27th April 2020) showed evidence of an overactive immune response in a handful of studies in cross-sectional analyses all done at a single time point.

*Added value of this study:* To determine the role of the immune response in a disease process, it is necessary to correlate immune activity with clinical parameters dynamically. In this study patients presented to hospital at different stages of disease so we took samples at different time-points to provide an accurate picture of the relevant pathobiology. In order to avoid loss of large components of the immune system due to the processes of storage, longitudinal samples were interrogated in real time to reveal the full immune alterations in COVID-19.

*Implications of all the available evidence:* Respiratory viruses continue to cause devastating global disease. The finding of altered myelopoiesis, with excess neutrophils and altered monocyte function, as dominant features in our study provides an incentive for clinical testing of therapeutics that specifically target this pathobiology. Given that inflammation is greatest prior to admission to intensive care, trials of specific immune-modulating therapies should be considered earlier in admission. Future studies of COVID-19 mechanisms should place more emphasis on longitudinal analyses since disease changes dramatically over time.

## Introduction

Severe acute respiratory syndrome coronavirus 2 (SARS-CoV-2) infection results in the clinical syndrome COVID-19 which, to date, has resulted in over 5 million confirmed cases and in excess of 330,000 attributable deaths world-wide. A large number of clinical trials have been established to evaluate anti-viral and immune modulatory strategies aimed at improving clinical outcome.

SARS-CoV-2 is a single stranded, positive sense RNA virus that enters cells via human angiotensin converting enzyme 2 (ACE2)^1^. Overlapping immune mechanisms exist to detect every stage of viral replication. Pattern recognition receptors of the innate immune system recognize viral antigen and virus-induced damage, increasing bone marrow hematopoiesis and immune cell mobilisation. If inflammatory mediator release is not controlled in duration and amplitude then “emergency hematopoiesis” leads to bystander tissue damage and the cytokine storm syndrome manifesting as organ dysfunction. Initial studies suggest cytokine storm syndrome occurs in COVID-19^2^. Indeed, neutrophilia and lymphopenia (resulting in an increased neutrophil:lymphocyte ratio), increased systemic interleukin-6 (IL-6) and C-reactive protein correlate with incidence of ICU admission and mortality^3^.

The Coronavirus Immune Response and Clinical Outcomes (CIRCO) study was conducted at four hospitals in Greater Manchester, UK. The aim was to identify demographic, clinical and immunological factors associated with different immune phenotypes in response to SARS-Cov-2 infection in hospitalised patients. Single time point analysis of immunity (e.g at admission) usually involves thousands of patients. However, we felt it was important to study the kinetics of the immune response at depth in a longitudinal well-defined patient cohort. Understanding the specific elements and kinetics of immune excess is critical to gain insight into immune phenotypes associated with disease progression, identify potential biomarkers that predict clinical outcomes and determine at which stage of the disease immune modulation may be effective^2^.

## Methods

### Cases

Between 29^th^ March and 7^th^ May, 2020, adults requiring hospital admission with suspected COVID-19 were recruited from 4 hospitals in the Greater Manchester area. Informed consent was obtained for each patient. Samples were collected at Manchester University Foundation Trust (MFT), Salford Royal NHS Foundation Trust (SRFT) and Pennine Acute NHS Trust (PAT) under the framework of the Manchester Allergy, Respiratory and Thoracic Surgery (ManARTS) Biobank (study no M2020-88) for MFT or the Northern Care Alliance Research Collection (NCARC) tissue biobank (study no NCA-009) for SRFT and PAT. Ethical approval obtained from the National Research Ethics Service (REC reference 15/NW/0409 for ManARTS and 18/WA/0368 for NCARC). Clinical information was extracted from written/electronic medical records including demographic data, presenting symptoms, comorbidities, radiographic findings, vital signs, and laboratory data (differential white blood cell count, CRP, kidney and liver function). Patients were included if they tested positive for SARS-CoV-2 by reverse-transcriptase–polymerase-chain-reaction (RT-PCR) on nasopharyngeal/oropharyngeal swabs or sputum. Patients with negative nasopharyngeal RT-PCR results were also included if there was a high clinical suspicion of COVID-19, the radiological findings supported the diagnosis and there was no other explanation for symptoms. Patients were excluded if an alternative diagnosis was reached, where indeterminate imaging findings were combined with negative SARS-CoV-2 nasopharyngeal (NP) test or there was another confounding acute illness not directly related to COVID-19. The severity of disease was scored each day, based on criteria for escalation of care (appendix 1). Patients were not stratified for disease severity if there was no available clinical observation data or patients were recruited more than 7 days after hospital admission. Where severity of disease changed during admission, the highest disease severity score was selected for classification. The first available time point was used for all cross-sectional comparisons between mild, moderate and severe disease. Peripheral blood samples were collected as soon after admission as possible and at 1-2 day intervals thereafter.

### Healthy

Recruiting healthy individuals from the community for blood sampling during the SARS-CoV-2 outbreak was not possible, and so we took three other approaches. The first was to sample frontline workers from Manchester University and NHS Trusts with an age range that matched our COVID-19 patients. Using this strategy peripheral blood samples were collected from 18 females and 9 males (age range 28-69). Secondly, we noted that most COVID-19 patients with mild disease were indistinguishable from our control group, which lent support to the validity of our comparisons. Finally, longitudinal analysis of patients with severe disease showed that they returned to our control normal range on recovery.

### Flow cytometry

White blood cells from lysed whole blood and peripheral blood mononuclear cells (PBMCs) separated by density gradient centrifugation were stained immediately on receipt with five panels of 20 antibodies (see appendix 2 for antibody clones and manufacturers) and immune cell profiles determined by flow cytometry. In order to assess the general immune responsiveness of immune cells, PBMCs were also stimulated *in vitro* for 3 hours with 10 ng/ml LPS in the presence of 10 ug/ml Brefeldin A to allow accumulation and analysis of intracellular proteins by flow cytometry. Cells were cultured in RPMI containing 10% fetal calf serum, L-Glutamine, Non-essential Amino Acids, HEPES and penicillin plus streptomycin (Gibco). Samples were acquired on an LSRFortessa cell analyzer (Becton Dickinson) and analysed using FlowJo (TreeStar).

### LEGENDplex

At multiple stages post-admission, 13 different mediators associated with anti-viral responses were measured in serum using LEGENDplex assays (BioLegend, San Diego, USA) according to the manufacturer’s instructions.

### Statistics

Results are presented as individual data points with medians. Groups were compared using an unpaired Mann-Whitney test for healthy individuals versus COVID-19 patients, Kruskal-Wallis test for multiple comparisons, or Spearman’s rank correlation coefficient test for correlation of separate parameters within the COVID-19 patient group, using Prism 8 software (GraphPad).

## Results

In total, 73 patients were recruited and 49 were stratified for disease severity. Six patients were excluded due to an alternative diagnosis (2 patients), indeterminate imaging findings with negative for SARS-CoV-2 NP test (2 patients) or a confounding acute illness was diagnosed (2 patients). Two patients could not be stratified for disease severity due to insufficient clinical observation data and a further 16 because recruitment occurred more than 7 days after admission. The median time from symptom onset to hospital admission was 7 days (IQR - 4-10). The overall median age was 61 (IQR 51-71) and 63% were male. The most frequent co-morbidities were diabetes, ischaemic heart disease, hypertension, asthma and chronic obstructive pulmonary disease (table 1). The majority (86%) of patients tested positive for SARS-CoV-2 via nasopharyngeal RT-PCR. In 14% of patients, symptoms and radiographic features were highly suggestive of COVID-19, but nasopharyngeal tested negative for the virus and thus a clinical diagnosis was made. Complications occurred in 8 out of 49 patients and included pulmonary embolism and acute kidney injury.

**Table 1.**
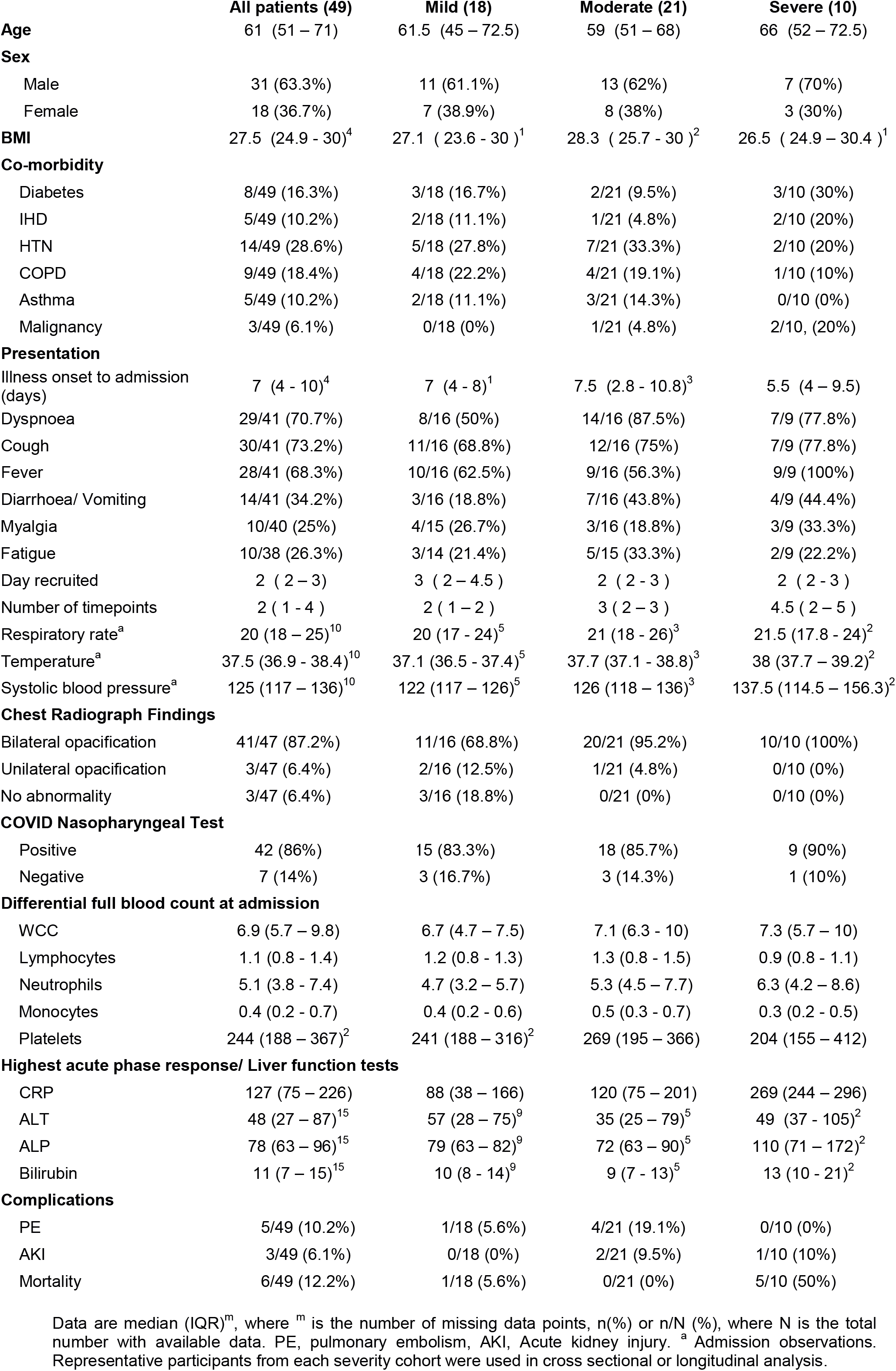

|  | All patients (49) | Mild (18) | Moderate (21) | Severe (10) |
| --- | --- | --- | --- | --- |
| **Age** | 61 (51 – 71) | 61.5 (45 – 72.5) | 59 (51 – 68) | 66 (52 – 72.5) |
| **Sex** |  |  |  |  |
| Male | 31 (63.3%) | 11 (61.1%) | 13 (62%) | 7 (70%) |
| Female | 18 (36.7%) | 7 (38.9%) | 8 (38%) | 3 (30%) |
| **BMI** | 27.5 (24.9 - 30)<sup>4</sup> | 27.1 (23.6 - 30)<sup>1</sup> | 28.3 (25.7 - 30)<sup>2</sup> | 26.5 (24.9 – 30.4)<sup>1</sup> |
| **Co-morbidity** |  |  |  |  |
| Diabetes | 8/49 (16.3%) | 3/18 (16.7%) | 2/21 (9.5%) | 3/10 (30%) |
| IHD | 5/49 (10.2%) | 2/18 (11.1%) | 1/21 (4.8%) | 2/10 (20%) |
| HTN | 14/49 (28.6%) | 5/18 (27.8%) | 7/21 (33.3%) | 2/10 (20%) |
| COPD | 9/49 (18.4%) | 4/18 (22.2%) | 4/21 (19.1%) | 1/10 (10%) |
| Asthma | 5/49 (10.2%) | 2/18 (11.1%) | 3/21 (14.3%) | 0/10 (0%) |
| Malignancy | 3/49 (6.1%) | 0/18 (0%) | 1/21 (4.8%) | 2/10, (20%) |
| **Presentation** |  |  |  |  |
| Illness onset to admission (days) | 7 (4 - 10)<sup>4</sup> | 7 (4 - 8)<sup>1</sup> | 7.5 (2.8 - 10.8)<sup>3</sup> | 5.5 (4 – 9.5) |
| Dyspnoea | 29/41 (70.7%) | 8/16 (50%) | 14/16 (87.5%) | 7/9 (77.8%) |
| Cough | 30/41 (73.2%) | 11/16 (68.8%) | 12/16 (75%) | 7/9 (77.8%) |
| Fever | 28/41 (68.3%) | 10/16 (62.5%) | 9/16 (56.3%) | 9/9 (100%) |
| Diarrhoea/ Vomiting | 14/41 (34.2%) | 3/16 (18.8%) | 7/16 (43.8%) | 4/9 (44.4%) |
| Myalgia | 10/40 (25%) | 4/15 (26.7%) | 3/16 (18.8%) | 3/9 (33.3%) |
| Fatigue | 10/38 (26.3%) | 3/14 (21.4%) | 5/15 (33.3%) | 2/9 (22.2%) |
| Day recruited | 2 (2 – 3) | 3 (2 – 4.5) | 2 (2 - 3) | 2 (2 - 3) |
| Number of timepoints | 2 (1 - 4) | 2 (1 – 2) | 3 (2 – 3) | 4.5 (2 – 5) |
| Respiratory rate<sup>a</sup> | 20 (18 – 25)<sup>10</sup> | 20 (17 - 24)<sup>5</sup> | 21 (18 - 26)<sup>3</sup> | 21.5 (17.8 - 24)<sup>2</sup> |
| Temperature<sup>a</sup> | 37.5 (36.9 - 38.4)<sup>10</sup> | 37.1 (36.5 - 37.4)<sup>5</sup> | 37.7 (37.1 - 38.8)<sup>3</sup> | 38 (37.7 – 39.2)<sup>2</sup> |
| Systolic blood pressure<sup>a</sup> | 125 (117 – 136)<sup>10</sup> | 122 (117 – 126)<sup>5</sup> | 126 (118 – 136)<sup>3</sup> | 137.5 (114.5 – 156.3)<sup>2</sup> |
| **Chest Radiograph Findings** |  |  |  |  |
| Bilateral opacification | 41/47 (87.2%) | 11/16 (68.8%) | 20/21 (95.2%) | 10/10 (100%) |
| Unilateral opacification | 3/47 (6.4%) | 2/16 (12.5%) | 1/21 (4.8%) | 0/10 (0%) |
| No abnormality | 3/47 (6.4%) | 3/16 (18.8%) | 0/21 (0%) | 0/10 (0%) |
| **COVID Nasopharyngeal Test** |  |  |  |  |
| Positive | 42 (86%) | 15 (83.3%) | 18 (85.7%) | 9 (90%) |
| Negative | 7 (14%) | 3 (16.7%) | 3 (14.3%) | 1 (10%) |
| **Differential full blood count at admission** |  |  |  |  |
| WCC | 6.9 (5.7 – 9.8) | 6.7 (4.7 – 7.5) | 7.1 (6.3 - 10) | 7.3 (5.7 – 10) |
| Lymphocytes | 1.1 (0.8 - 1.4) | 1.2 (0.8 - 1.3) | 1.3 (0.8 - 1.5) | 0.9 (0.8 - 1.1) |
| Neutrophils | 5.1 (3.8 - 7.4) | 4.7 (3.2 – 5.7) | 5.3 (4.5 – 7.7) | 6.3 (4.2 – 8.6) |
| Monocytes | 0.4 (0.2 - 0.7) | 0.4 (0.2 - 0.6) | 0.5 (0.3 - 0.7) | 0.3 (0.2 - 0.5) |
| Platelets | 244 (188 – 367)<sup>2</sup> | 241 (188 – 316)<sup>2</sup> | 269 (195 – 366) | 204 (155 – 412) |
| **Highest acute phase response/ Liver function tests** |  |  |  |  |
| CRP | 127 (75 – 226) | 88 (38 – 166) | 120 (75 – 201) | 269 (244 – 296) |
| ALT | 48 (27 – 87)<sup>15</sup> | 57 (28 – 75)<sup>9</sup> | 35 (25 – 79)<sup>5</sup> | 49 (37 - 105)<sup>2</sup> |
| ALP | 78 (63 – 96)<sup>15</sup> | 79 (63 – 82)<sup>9</sup> | 72 (63 – 90)<sup>5</sup> | 110 (71 – 172)<sup>2</sup> |
| Bilirubin | 11 (7 – 15)<sup>15</sup> | 10 (8 - 14)<sup>9</sup> | 9 (7 - 13)<sup>5</sup> | 13 (10 - 21)<sup>2</sup> |
| **Complications** |  |  |  |  |
| PE | 5/49 (10.2%) | 1/18 (5.6%) | 4/21 (19.1%) | 0/10 (0%) |
| AKI | 3/49 (6.1%) | 0/18 (0%) | 2/21 (9.5%) | 1/10 (10%) |
| Mortality | 6/49 (12.2%) | 1/18 (5.6%) | 0/21 (0%) | 5/10 (50%) |

Data are median (IQR)<sup>m</sup>, where <sup>m</sup> is the number of missing data points, n(%) or n/N (%), where N is the total number with available data. PE, pulmonary embolism, AKI, Acute kidney injury. <sup>a</sup> Admission observations. Representative participants from each severity cohort were used in cross sectional or longitudinal analysis.

Patient disease severity was defined as mild (less than 28% FiO2), moderate (28-60% FiO2) or severe (above 60% FiO2, or admission to intensive care) (appendix 1). All severe patients were recruited from ward level care. Only 1 patient met the criteria for direct ICU admission. Out of the 10 severe patients, 7 were treated in intensive care, 4 were intubated and ventilated and 2 were treated by CPAP alone (data not available for 1 patient). Three severe patients had ceilings of care in place that specified ward level care due to frailty. Death occurred in 50% of severe cases of COVID-19.

On hospital admission little difference was observed in total white blood cell counts, neutrophils or lymphocytes (figure 1A). Although, as reported previously^4^, a higher neutrophil to lymphocyte ratio (NLR) on hospital admission was observed in those patients whose disease trajectory was ultimately severe, whereas there were no appreciable differences observed in monocytes (figure 1A, 1B and table 1). By only analysing differential white cell counts, important correlations may be missed. We, therefore, performed multidimensional flow cytometry covering over 80 immune markers across 5 panels on whole blood cells and separated PBMCs.

**Figure 1.**
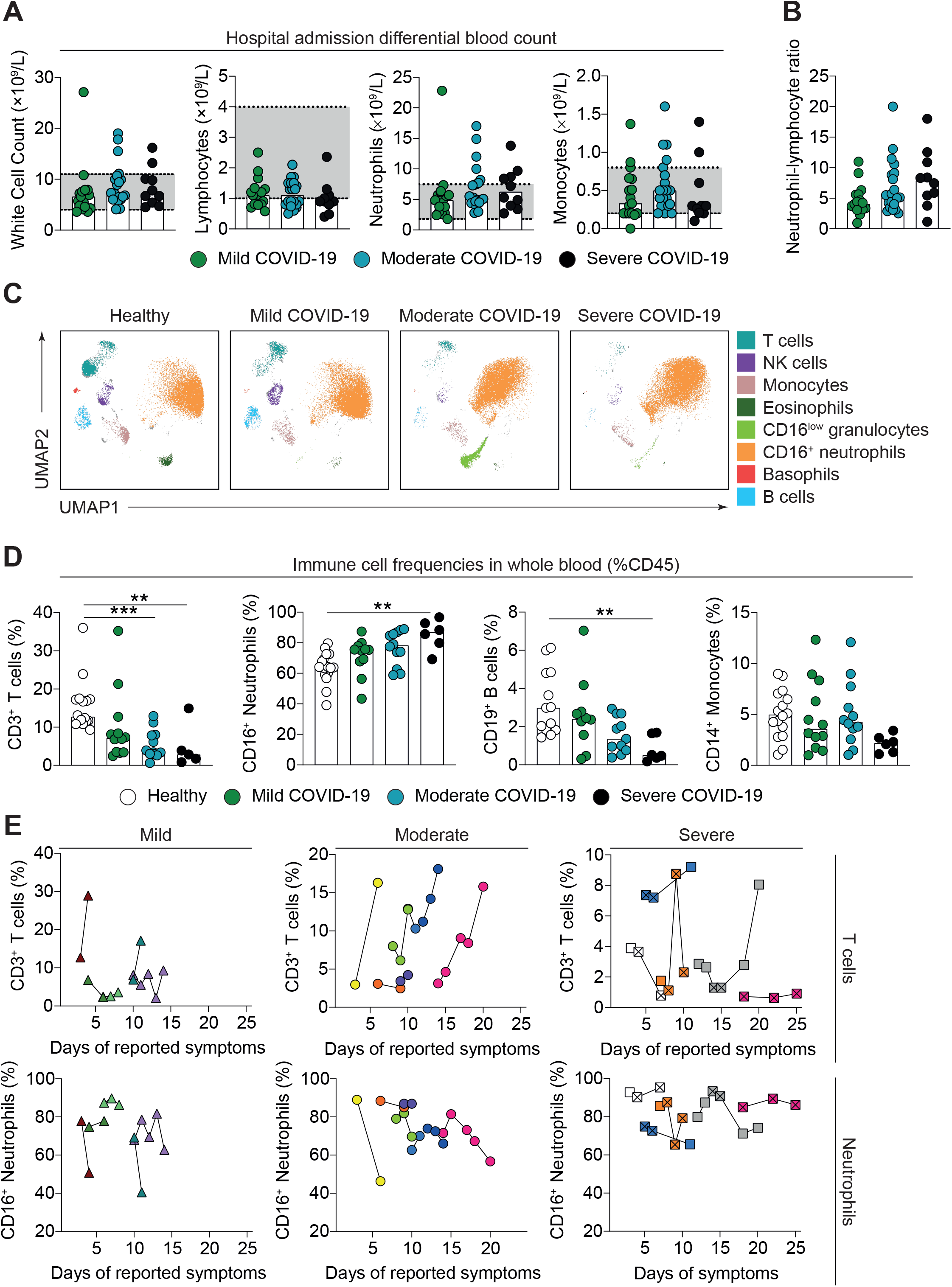
Whole blood immune profile of COVID-19 patients. (**A**) Hospital assessed white blood cell count (WCC), lymphocyte count, monocyte count, neutrophil count and (**B**) Neutrophil to lymphocyte ratio (NLR) calculated using hospital assessed neutrophil and lymphocyte counts. Grey region represents normal range. Patients stratified into mild (n=17), moderate (n=20) and severe (n=10) disease groups. (**C**) Uniform Manifold Approximation and Projection (UMAP) of flow cytometry panel broadly visualising white cells in whole blood. Representative images for healthy individuals, mild, moderate and severe patients are shown. (**D**) Neutrophil, monocyte and lymphocyte frequencies for healthy individuals and recruitment samples for COVID-19 patients, according to disease severity. Mild (n=12), moderate (n=13) and severe (n=5). (**E**) Longitudinal time course of CD3^+^ T cells and CD16^+^ neutrophils segregated by disease severity. Individual patients are shown as different colours and shapes. Crossed squares for severe patients are time points in ITU. For comparisons of disease subtypes and healthy individuals: Kruskal-Wallis test with multiple comparisons was carried out. (*P<0.05, **P<0.01, ***P<0.001).

During a pandemic, it is impossible to recruit healthy individuals for blood sampling from the community. As the data emerged, it became clear that the patients in the mild disease category were indistinguishable in the majority of analyses from our frontline volunteers. Furthermore, comparing the results across the different disease severities provided additional validation of our control group that tracked most closely with the patients experiencing mild disease. Typically, all parameters returned to within control levels on recovery, irrespective of disease severity.

Using Uniform Manifold Approximation and Projection (UMAP) we were able to visualise global immune cell differences between COVID-19 patients and healthy individuals (figure 1C). Broadly, this included alterations in the characteristics of neutrophils and monocytes and decreases in B cells, T cells, basophils and NK cells in the initial recruitment blood sample, taken within 7 days of hospital admission (figure 1D and appendix 3). These differences were exaggerated in patients with severe disease. This global picture was confirmed by manual flow cytometric gating (figure 1D). In addition, the frequency of basophils in whole blood and plasmacytoid dendritic cells (pDCs) within PBMCs were significantly decreased in patients compared to healthy individuals (appendix 3A-B). Therefore, hospital differential count (on day of admission) and laboratory (on day of recruitment to study) analysis of overall immune cell populations concur. Longitudinal analysis revealed that in the majority of patients (70%) (irrespective of severity) T cell frequencies in whole blood increased prior to hospital discharge, while neutrophil frequencies reciprocally decreased (figure 1E). In two severe patients who did not recover, T cell frequencies were extremely low and neutrophil frequencies high (figure 1E, white and pink squares). This suggests that recovery is associated with a reduction of neutrophils and an increase in T cells.

Within the reduced T cell population there were no dramatic alterations in CD4+ or CD8+ T cell frequencies, other than a slight decrease in CD4+ T cells in severe COVID-19 patients (figure 2A and 2B). Both T cell subsets showed signs of activation in COVID-19 patients; however, this was more striking in CD8+ T cells, did not track with disease severity, and was highly variable amongst patients (appendix 4A-C). No differences were observed in the expression of the regulatory molecule PD-1 (appendix 4D). Interestingly, in 34/43 COVID-19 patients higher perforin expression was observed in CD8+ T cells compared to healthy individuals (appendix figure 4E), implying these cells have activated a cytotoxic programme. Perforin expression in CD8+ T cells did not significantly track with disease severity (figure 2C,D), although we did observe a correlation with CRP (figure 2E). Therefore, only minor differences were observed in the CD8+ T cell compartment.

**Figure 2.**
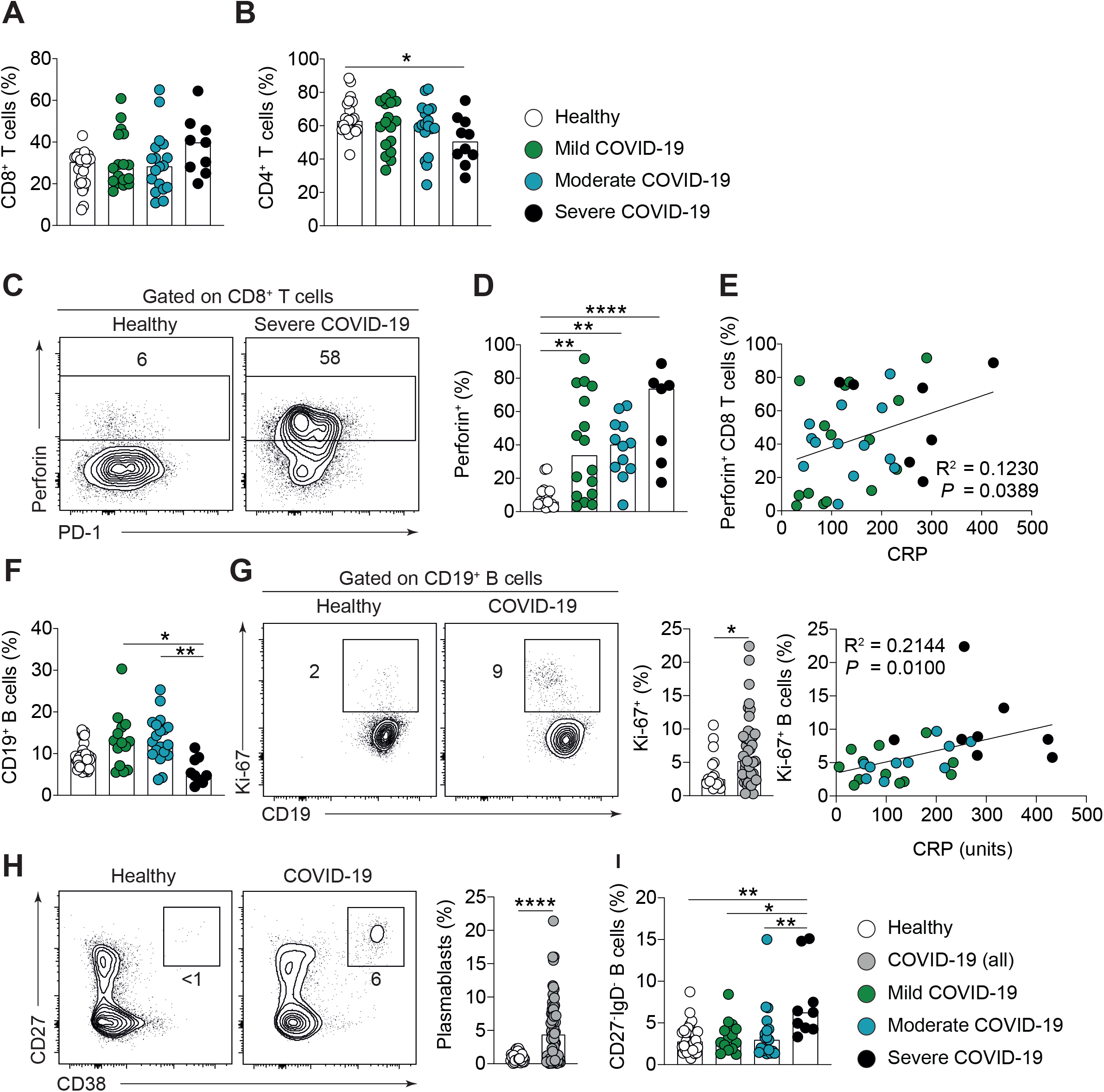
Altered phenotype of T and B cells in COVID-19 patients. **(A**,**B)** Cumulative data show frequency of (**A**) CD8^+^ and (**B**) CD4^+^ T cells in PBMC of healthy individuals (n=24) and COVID-19 patients with mild (n=17), moderate (n=18) and severe (n=9-10) disease. **(C**,**D)** Representative flow cytometry plots and cumulative data show frequencies of CD8^+^ T cells which are positive for Perforin in healthy individuals (n=16) and COVID-19 patients with mild (n=16), moderate (n=12) and severe (n=7) disease. **(E)** Graph showing correlation of Perforin^+^ CD8^+^ T cell frequency with C-reactive protein (CRP) in COVID-19 patients. **(F)** Cumulative data show *ex vivo* frequency of CD19^+^ B cells in healthy individuals (n=31) and COVID-19 patients with mild (n=14), moderate (n=19) and severe (n=9) disease. **(G)** Representative flow cytometry plots and cumulative data show Ki67 expression by B cells in healthy individuals (n=16) and COVID-19 patients (n=45). Cumulative graph shows correlation of Ki67^+^B cells with C-reactive protein (CRP). **(H)** Representative flow cytometry plots and cumulative data show frequency of CD27^hi^CD38^hi^ plasmablasts in healthy individuals (n=31) and COVID-19 patients (n=66). **(I)** Cumulative data show frequency of double negative (CD27^-^IgD^-^) B cells in healthy individuals (n=31) and COVID-19 patients with mild (n=14), moderate (n=19) and severe (n=9) disease. For healthy individuals versus COVID-19: Mann-Whitney test was carried out. For comparisons of disease subtypes and healthy individuals: Kruskal-Wallis test with multiple comparisons was carried out. For correlations: Spearman ranked coefficient correlation test was carried out (*P<0.05; **P<0.01; P<0.001; ns, not significant).

The frequency of B cells was also reduced in COVID-19 patients with severe disease compared to patients with mild and moderate disease (figure 2F). Although reduced, B cells displayed an increased expression of Ki67 (indicative of activation), which positively correlated with CRP levels (figure 2G). When examining B cell subsets no major differences in naive and switched memory B cell subsets were observed in COVID-19 patients (appendix 5A and 5B). However, we observed an expansion of antibody-secreting plasmablasts (CD27^hi^CD38^hi^CD24^-^) in COVID-19 patients, which did not track with disease severity (figure 2H and appendix 5C). The only other population of B cells raised in patients with severe COVID-19 compared to patients with mild and moderate disease was a significant expansion of double negative (DN) B cells (CD27-IgD-) (figure 2I and appendix 5A). DN B cells have previously been shown to exhibit an exhausted phenotype in patients with HIV. This suggests that patients with severe COVID-19 may generate less effective B cell responses.

By far the most significant changes were observed in monocytes. In mild COVID-19 there was an expansion of the inflammatory CD14^+^CD16^+^ (intermediate) sub-population (appendix 6A), and enhanced CD64, (high affinity Fc receptor for monomeric IgG (Fc*γ*RI)) on CD14^+^ classical monocytes (figure 3A). Upon stimulation with LPS, monocytes from mild COVID-19 also displayed increased intracellular TNF, which significantly correlated with CD64 (figure 3B, C) whilst IL-1*β* levels were lower in monocytes from COVID-19 patients (appendix 6D). Severe COVID-19, on the other hand, was associated with monocytes displaying increased expression of the cell cycle marker, Ki67 (normally <5% in healthy peripheral blood), irrespective of whether monocytes were stimulated or not (figure 3C and appendix 6C), which strongly correlated with hospital data for CRP (figure 3C). We next examined cyclooxygenase-2 (COX-2) expression in monocytes, which is an enzyme involved in the production of prostaglandins. Remarkably, an early reduction of COX-2 was evident in patients that went on to develop severe COVID-19 (figure 3D).

**Figure 3.**
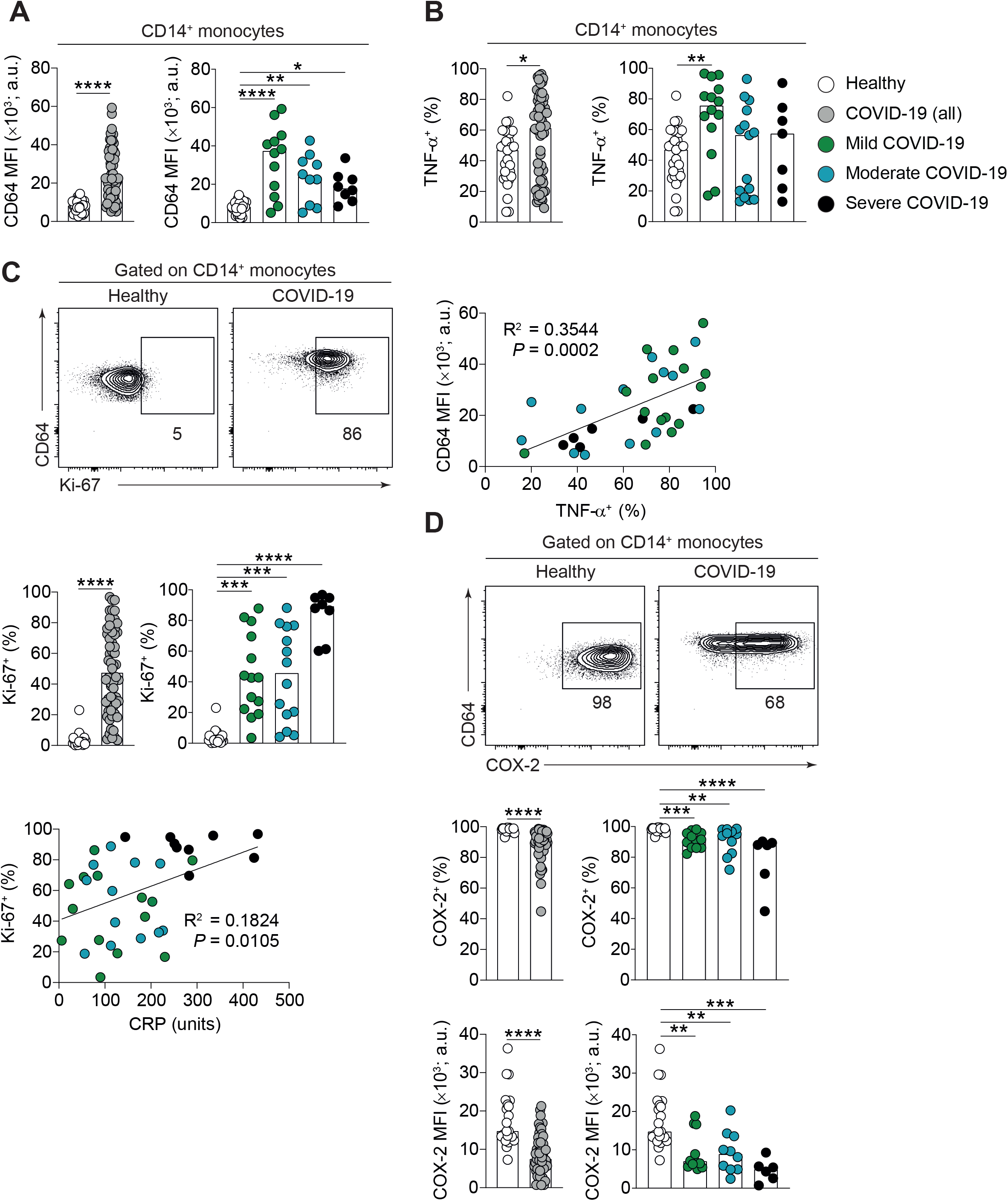
Dysregulation of circulating monocytes in COVID-19. **(A)** Levels of CD64 as assessed by mean fluorescence intensity (MFI) on CD14^+^classical monocytes in healthy individuals (n=25) and total COVID-19 patients (n=58). COVID-19 patients were stratified into mild (n=12), moderate (n=10) and severe (n=8) disease. **(B)** Production of TNFα by CD14^+^ monocytes following LPS stimulation and correlation of CD64 levels (MFI) with TNFa production, in healthy individuals (n=29) and total COVID-19 patients (n=59). COVID-19 patients were stratified into mild (n=14), moderate (n=15) and severe (n=7) disease. **(C)** Representative FACS plots demonstrating intracellular Ki67 staining by CD14^+^monocytes, summary graphs demonstrating expression of Ki67 following LPS stimulation and correlation of Ki67 (% of monocytes expressing Ki67) with C-reactive protein (CRP) in healthy individuals (n=25) and total COVID-19 patients (n=60). COVID-19 patients were stratified into mild (n=14), moderate (n=14) and severe (n=8) disease. **(D)** Representative FACS plots demonstrating intracellular COX2 expression by CD14^+^monocytes and summary graphs demonstrating proportion of CD14^+^ monocytes expressing COX2 following LPS stimulation, and levels of expression of COX2 as assessed by MFI, in healthy individuals (n=21) and total COVID-19 patients (n=51). COVID-19 patients were stratified into mild (n=12), moderate (n=11) and severe (n=6) disease. Individual data points shown with median for healthy versus COVID-19 and disease subtypes. For healthy versus COVID-19: Mann-Whitney test was carried out. For comparisons of disease subtypes and healthy individuals: Kruskal-Wallis test with multiple comparisons was carried out. For correlations: Spearman ranked coefficient correlation test was carried out (*P<0.05; **P<0.01; P<0.001).

We next examined sequential samples per patient and plotted the results based on the time since reported onset of symptoms. CD8+ T cells remained remarkably stable over the disease course (appendix 7A and 7B). However, we noticed that in mild and moderate COVID-19 perforin expressing CD8+ T cells increased with time, with the highest level immediately prior to discharge (appendix 7C,D). This suggests development of functional CD8+ T cells is beneficial in COVID-19. Reduced B cells was associated with a severe disease trajectory (notably in one of the deceased patients) (appendix 7E), particularly the switched memory subset (appendix 7F). Collectively, this suggests that low B cell responses associate with severe disease. This reduction may be due to exhaustion that is supported by an expansion of double negative B cells in severe patients (appendix 7G). The most striking trajectories that correlated with development of severe COVID-19 were again in the monocyte population, where low COX-2 was present throughout (figure 4A) and higher Ki67 was observed prior to ITU admission (figure 4B). TNF expression following monocyte stimulation remained higher in patients with mild disease throughout the disease course (figure 4C).

**Figure 4.**
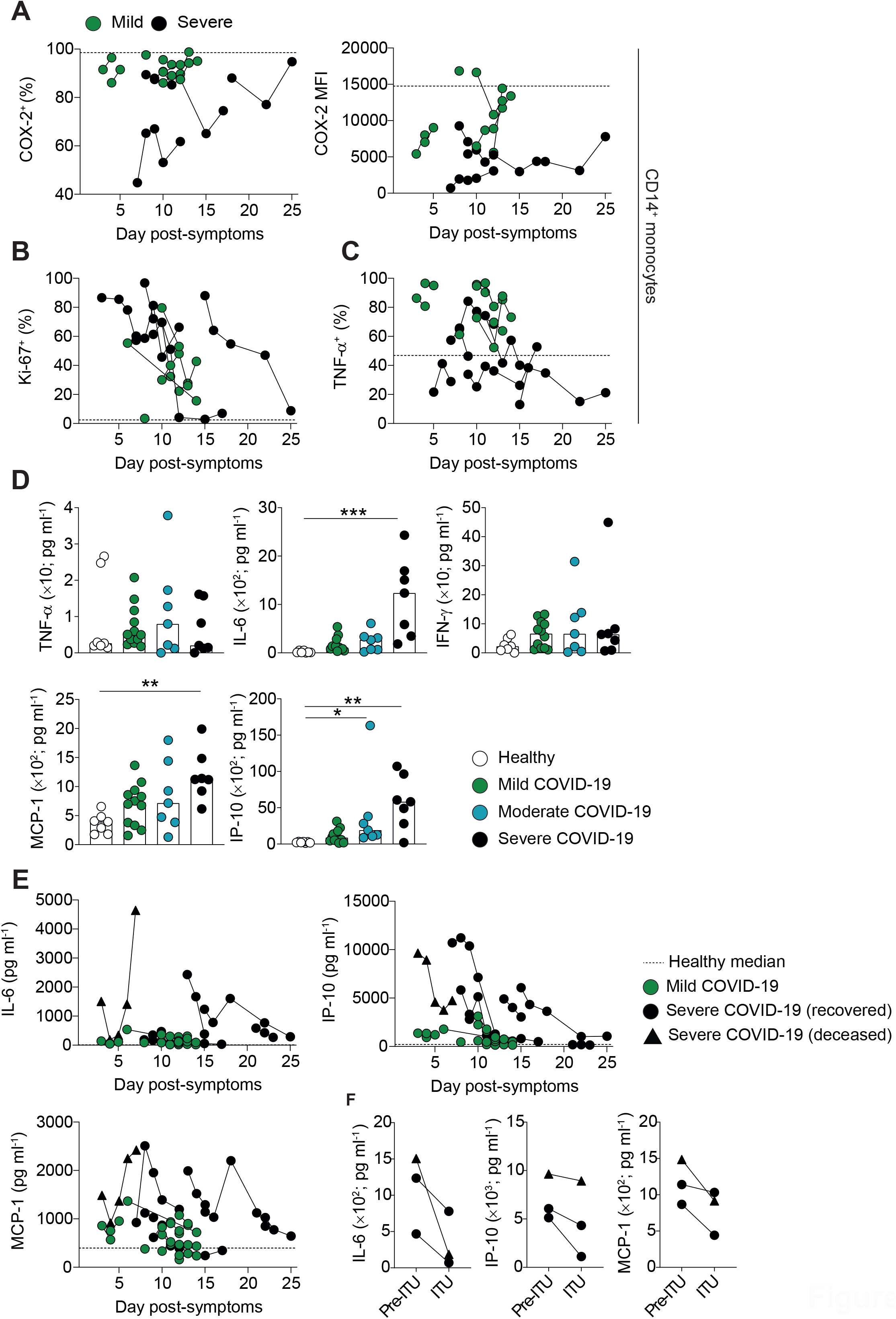
Longitudinal assessment of immune parameters during hospital admission. **(A)** Graph shows frequencies of CD14^+^ monocytes expressing COX2 following LPS stimulation, and levels of expression of COX2 as assessed by MFI in admitted patients over all measured time points, in mild (grey shapes, n=7), and severe (red shapes, n=4) COVID-19 patients. **(B)** Graph shows frequencies of CD14^+^ monocytes expressing Ki67 following LPS stimulation in admitted patients over all measured time points, in mild (grey shapes, n=6), and severe (red shapes, n=5) COVID-19 patients. **(C)** Graph shows frequencies of CD14^+^ monocytes producing TNFa following LPS stimulation in admitted patients over all measured time points, in mild (grey shapes, n=7) and severe (red shapes, n=6) COVID-19 patients. On all graphs x axis values represent the number of days since onset of symptoms and the dotted line the median value from healthy individuals. **(D)** Levels of systemic IL-6, MCP-1, IP-10, TNFα, and IFNγ were measured in serum from healthy individuals (n=7) and COVID-19 patients with mild (n=12), moderate (n=7) and severe (n=7) disease using LEGENDplex assay. **(E)** Graphs show levels of systemic IL-6, MCP-1 and IP-10 in admitted patients over all measured time points, in mild (green shapes; n=12) and severe (red shapes; n=7) COVID-19 patients. Deceased patient represented by triangles. On all graphs x axis values represent the number of days since reported onset of symptoms and the dotted line the median value from healthy individuals. (**F**) Serum levels of IL-6, IP-10 and MCP-1 at last time-point Pre-ITU and first time-point after transfer to ITU. For comparisons of disease subtypes and healthy individuals: Kruskal-Wallis test with multiple comparisons was carried out. (*P<0.05; **P<0.01; P<0.001).

We next assessed soluble inflammatory mediators on admission to hospital and across the length of hospital stay. Of the many mediators analysed in serum the ones significantly different between patients and healthy controls were IL-6, IL-10, monocyte-chemoattractant protein-1 (MCP-1) and interferon gamma-induced protein 10 (IP-10) (figure 4D and appendix 8). The increased levels of these cytokines were specific to COVID-19 patients with severe disease, and strongly indicate a role for myeloid cells in disease pathogenesis of COVID-19. Longitudinal analysis revealed that the highest levels of inflammatory mediators occurred early in disease trajectory following hospital admission, and critically, declined rapidly following admission to ITU despite no indications of recovery (figure 4E and 4F).

## Discussion

This detailed longitudinal analysis of COVID-19 patients of varying severity and outcome, the first reported here, has revealed common immunological features and aspects that track with severity. We provide information concerning pathogenesis that should influence clinical trials and therapeutics. An aspect not expected included the presence of proliferating (Ki67^+^) monocytes in the blood, with low COX-2 and high Fc receptor *γ* (FcRI, CD64). These properties all reverted in patients with good outcome. Also, not anticipated, was that multiple aspects of inflammation were diminished as patients progressed in severity and time. Thus, production of IL-6, IP-10, MCP-1 typically reduced late in disease, even in patients who did not recover.

The cellular analysis indicates considerable abnormalities in the myeloid compartment, much more so than the lymphopenia reported previously^5^. In the myeloid compartment, the profound neutrophilia, which reduces as patients improve, suggests that neutrophils and their products may contribute to the pathology. Activated neutrophils produce large quantities of S100A1 and A2, HMGB1 and free radicals. Furthermore, neutrophils release fibrous scaffolds containing DNA, histones, neutrophil elastase and myeloperoxidase that trap and kill pathogens, but also contribute to the development of acute respiratory distress syndrome^6,7^. To support their role, elevated neutrophil products have been identified in the sera of COVID-19 patients that correlate with clinical parameters such as C-reactive protein, D-dimer, and lactate dehydrogenase^8^.

The most unusual cell lineage in COVID-19 is the monocytes. In health, monocytes proliferate in the bone marrow and do not proliferate in blood, and may do so again in tissues as they differentiate^9^. Monocytes expressing proliferation markers in the blood have not previously been reported in COVID-19 patients. In these patients, up to 80% of monocytes were expressing proliferation marker Ki67, the highest consistently in patients with severe disease. The most likely explanation for this phenomenon is premature release of monocytes from the marrow, due to inflammatory signals. This has been previously reported in pandemic H1N1 influenza^10,11^.

Equally remarkable was the reduced expression of COX-2 in monocytes, from an early stage in disease in patients with a severe disease trajectory. COX-2 facilitates the production of prostanoids including prostaglandin E2 (PGE2) and increases lipid mediators involved in vasodilation, vascular permeability and immune cell chemotaxis^12^. It may also hinder the production of anti-inflammatory/pro-resolution prostanoids, the resolvins^13^. Some viruses (e.g avian H1N1 influenza the 2003 SARS coronavirus^14^) up-regulate the cyclo-oxygenase pathway to regulate viral replication. However, its reduction in monocytes in severe lung viral infection has not been reported previously. Reduced COX-2 alongside high IP-10, as seen here in severe COVID-19 patients, is an immune profile strongly associated with pathology in idiopathic pulmonary fibrosis (IPF)^15^. Therefore our data indicate a strong fibrotic signature in patients with severe disease, supporting studies observing an unusual pattern of fibrosis in COVID-19.

The function of monocytes in COVID-19 was also assessed by cytokine immunostaining. It was noteworthy that intracellular TNF was augmented in COVID-19 monocytes at hospital admission, supporting the hypothesis that early anti-TNF should be tested in COVID-19^16^.

Our data concurs with several features of the disease in Wuhan, China, and is corroborated in single cell RNA sequencing of bronchoalveolar lavage cells at a single time point^17^, but importantly we extend this into a longitudinal analysis that gives greater insight. Similarities include elevated CRP and IL-6 in patients at the time of hospitalization who eventually died^18^ and increased IP-10 (CXCL-10) in those who later developed severe disease^19^. IP-10 is an interferon-inducible chemokine receptor that facilitates directed migration of many immune cells^20^ and is elevated in other coronavirus infections including MERS-CoV and SARS-CoV^21^ and in Influenza virus of swine origin (H1N1)^22,23^. The heightened levels of monocyte-chemoattractant protein 1 (MCP-1) upon admission further indicate dysregulation of monocyte function and migration in patients with severe disease. Importantly, IL-6, IP-10 and MCP-1 levels are highest around the time of hospital admission, but are reduced rapidly as patients are admitted to ITU, which likely signifies exhaustion of immune cells and the lineages producing them.

There are, of course, limitations to any study of samples during a viral pandemic for which there is no vaccine. However, we believe that these do not reduce the importance of the messages derived from our study. A longitudinal analysis in real time for phenotypic, functional and soluble markers naturally limits the number of patients interrogated. In-depth analysis of smaller cohorts however, is necessary to gain insight into mechanism and is of interest to the pharmaceutical industry. In addition to the limitations of control subjects addressed above, the only other is that patients may not accurately define the onset of symptoms. As data is plotted per patient, however, this does not affect the interpretation of the results.

There are clinical implications of our data, as far as it has been analysed. Using non-steroidal anti-inflammatory drugs (NSAIDs) remains controversial^24^ and our study would suggest it is not desirable, as it would compound the already low COX-2^25^. Since most of the pathogenic mechanisms involve myeloid cells, neutrophils and monocytes, it would be advantageous to reduce their influx to the lung once lung pathology is established. Relevant strategies include inhibition of the complement anaphylatoxin C5a^26^ or IL-8 (CXCL8), which are strong chemoattractants for many immune cells, including neutrophils^27^. Antagonism of CXCR2 that mobilises neutrophil and monocyte from the bone marrow, neutrophil elastase inhibitors and inhibition of G-CSF, IL-23 and IL-17 that promote neutrophil survival, are also options^28^. Anti-IL-6 and anti-TNF agents are already being investigated for COVID-19 treatment and are relevant to neutrophils, which express the requisite cytokine receptors. Furthermore, JAK inhibitors are currently in clinical trials and may also reduce neutrophil levels^29^. Targeting toxic products of neutrophils such as S100A1/A2, HMGB1 and free radicals^30^, but also the formation of NETS could be beneficial^30^.

Finally, one of the clearest messages is that treating patients early after hospitalization is likely to be most beneficial - cytokines and cell levels are high and there is the potential for recovery. There are also potential predictors of severity which could stratify patients for treatment at admission, including high neutrophil/Tcell ratio, high Ki67 and low COX2 in monocytes.

## Data Availability

Individual participant data that underlie the results reported will be shared immediately following publication to anyone for any purpose. Data is available by contacting TH or JG and requestors will be required to sign a data access agreement.

## Acknowledgements

This report is independent research supported by the North West Lung Centre Charity and National Institute for Health Research Clinical Research Facility at Manchester University NHS Foundation Trust. The views expressed in this publication are those of the author(s) and not necessarily those of the NHS, the North West Lung Centre Charity, National Institute for Health Research or the Department of Health. The authors would like to acknowledge the Manchester Allergy, Respiratory and Thoracic Surgery Biobank, the Northern Care Alliance Research Collection tissue bank and the North West Lung Centre Charity for supporting this project. In addition, we would like to thank the Immunology community within the Lydia Becker Institute of Immunology and Inflammation, the core flow cytometry facility at the University of Manchester, the Manchester COVID-19 Rapid Response Group and the study participants for their contribution.

## Contributors

ERM, MM, SBK, JEK, CJ, TS, JRG and TH performed the study design, tissue processing, data collection, data analysis, data interpretation and manuscript writing.

SK and MR performed data analysis and generated the figures.

AU, NDB, PD, AS, TF performed sample collection and biobanking, AU, AS, TF contributed to data interpretation, presentation of clinical data and paper construction L-PH, RTRC, GL, CIRCO and MF contributed to study design, patient consent and sample collection and manuscript preparation

---

**Appendix Figure 1.**
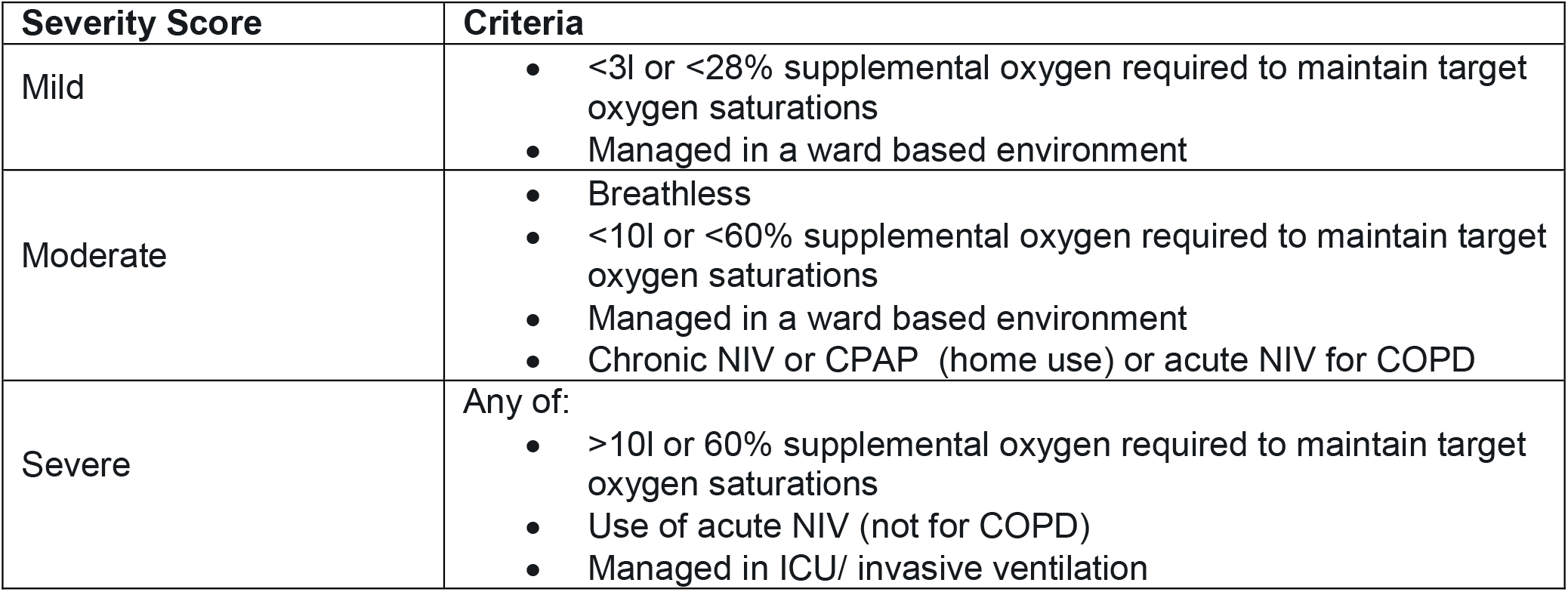

**Appendix Figure 2.**
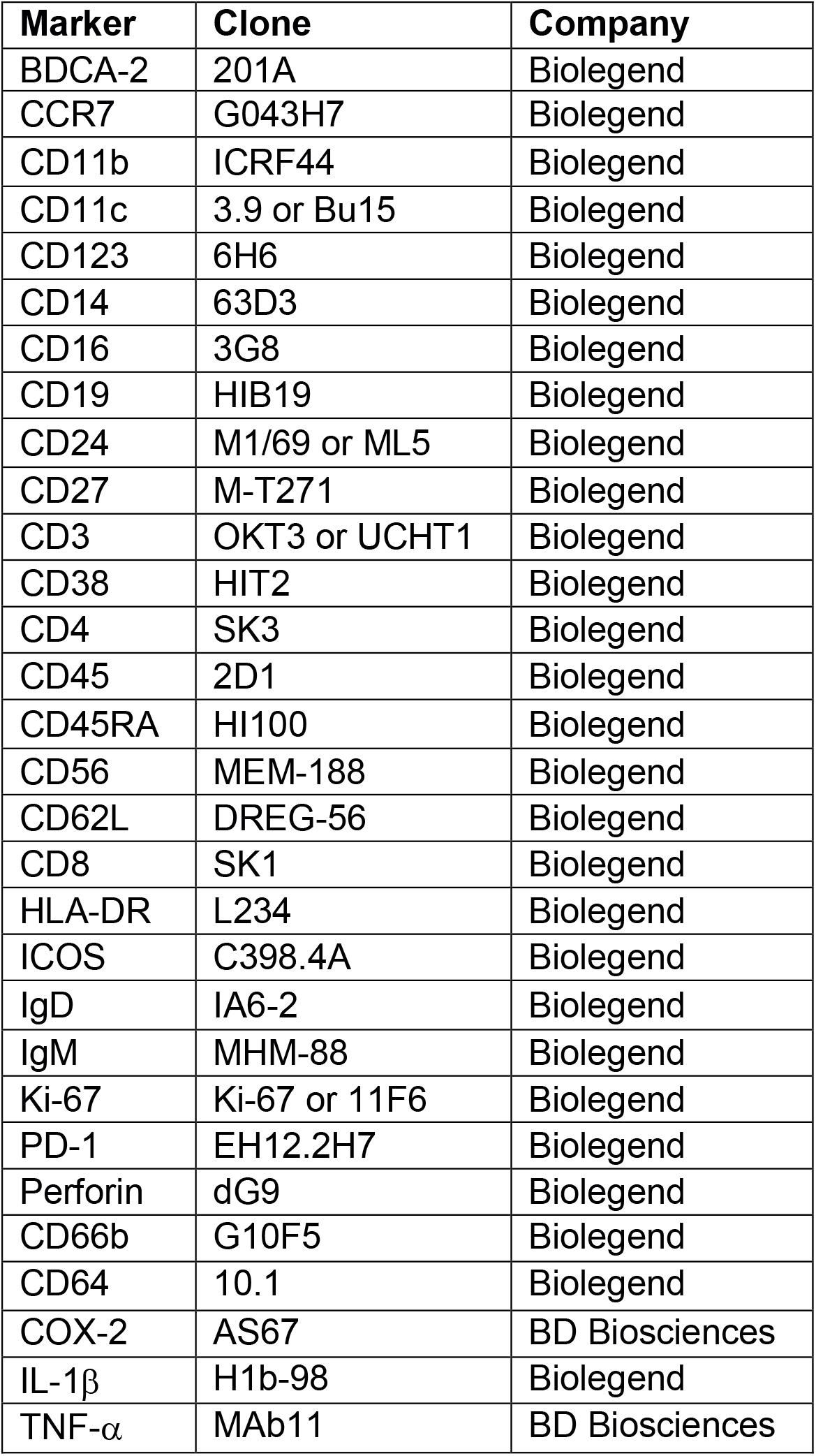

**Appendix Figure 3.**
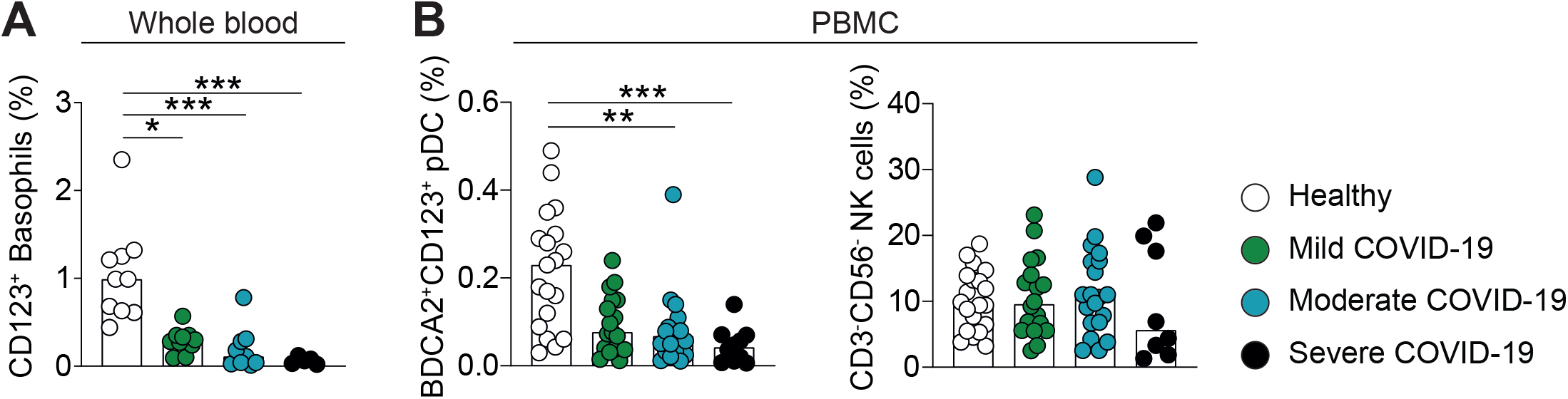

**Appendix Figure 4.**
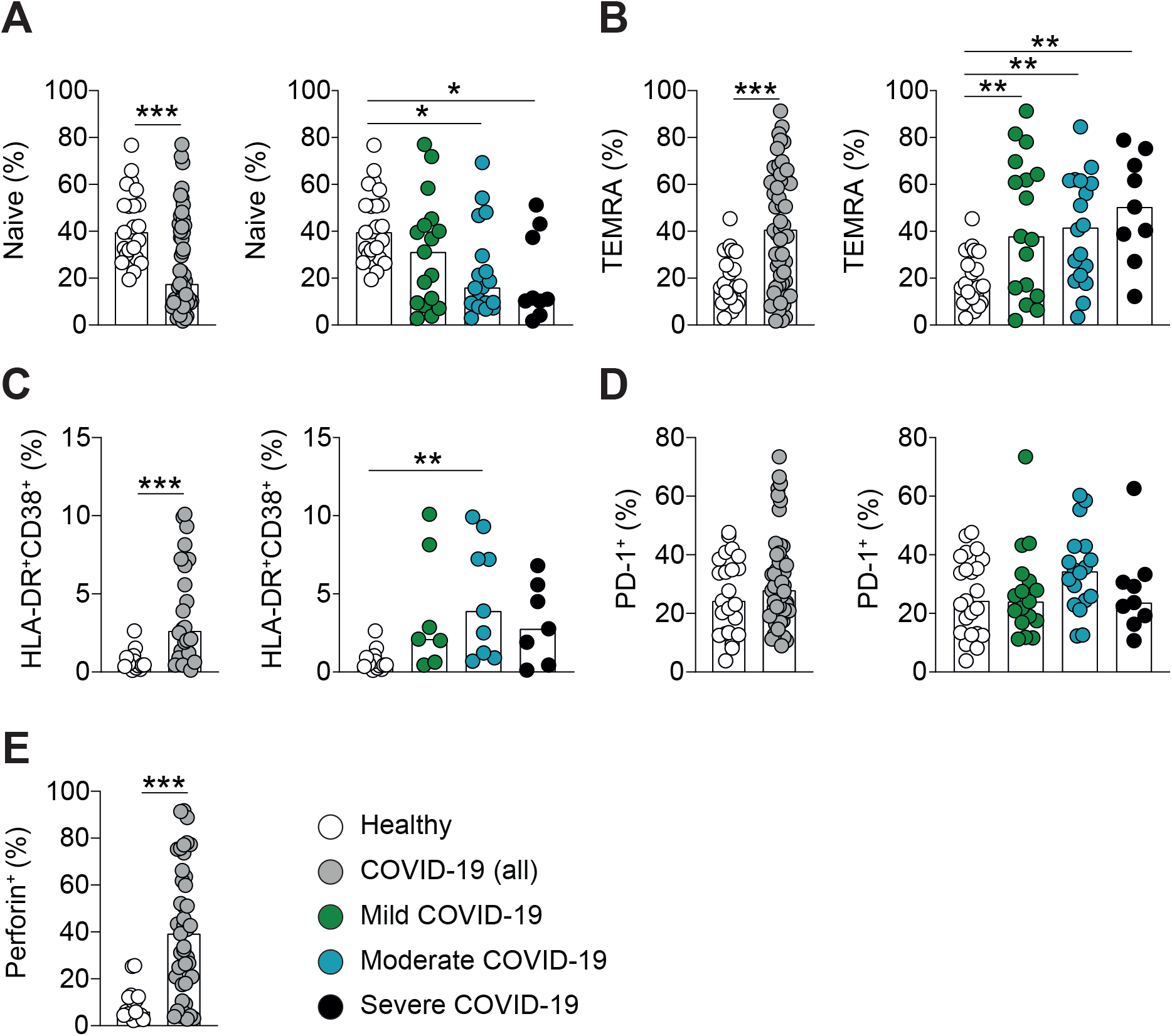

**Appendix Figure 5.**
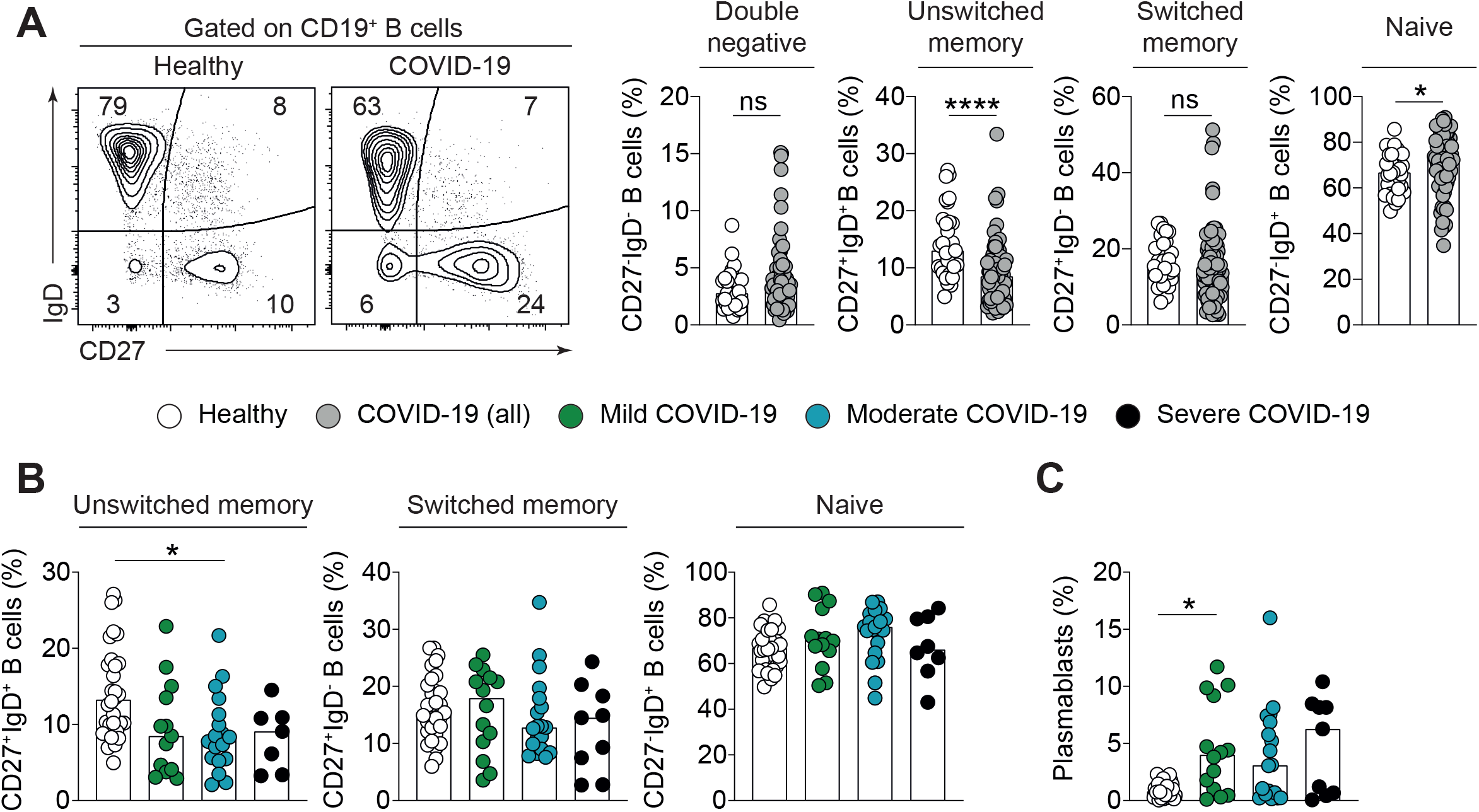

**Appendix Figure 6.**
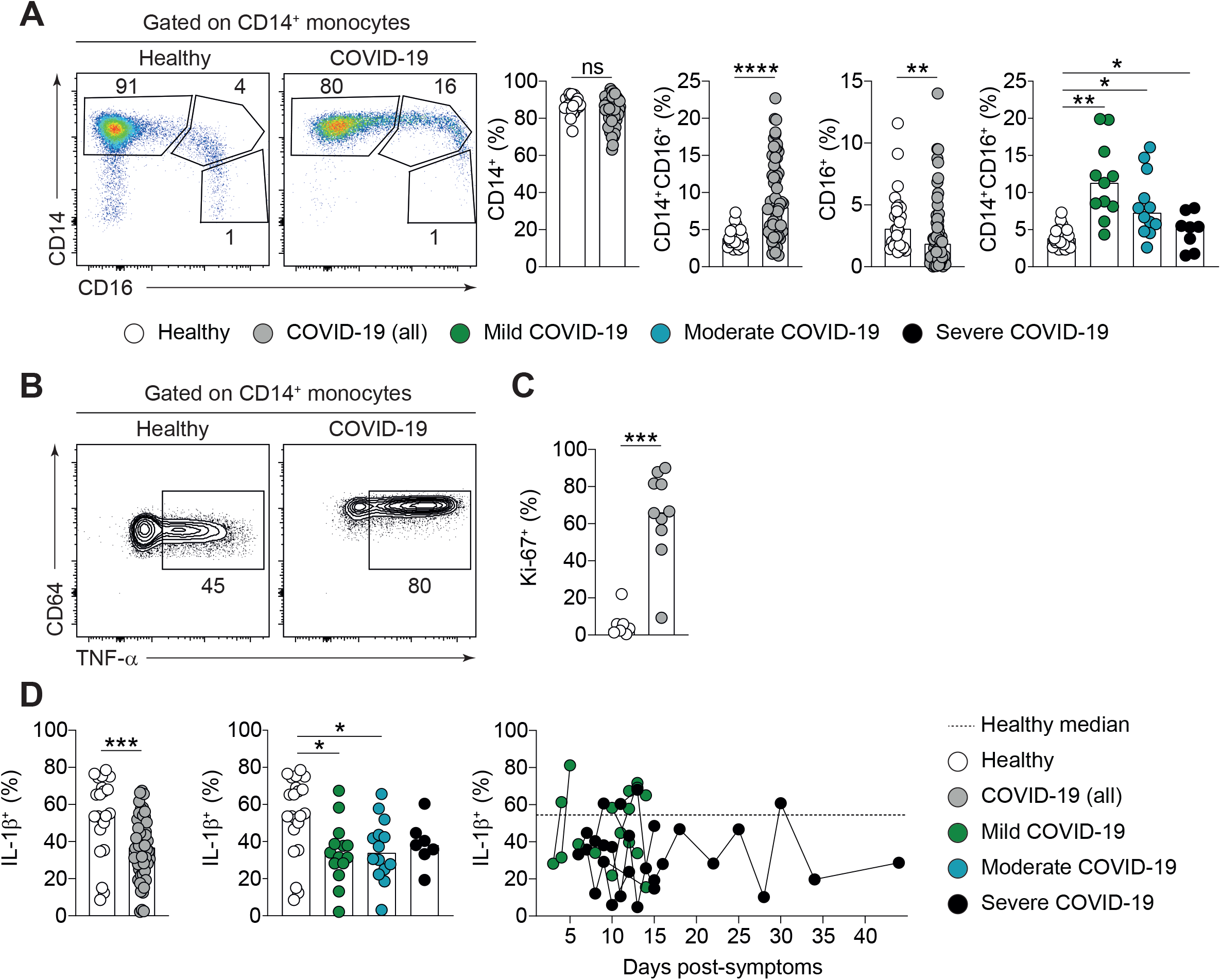

**Appendix Figure 7.**
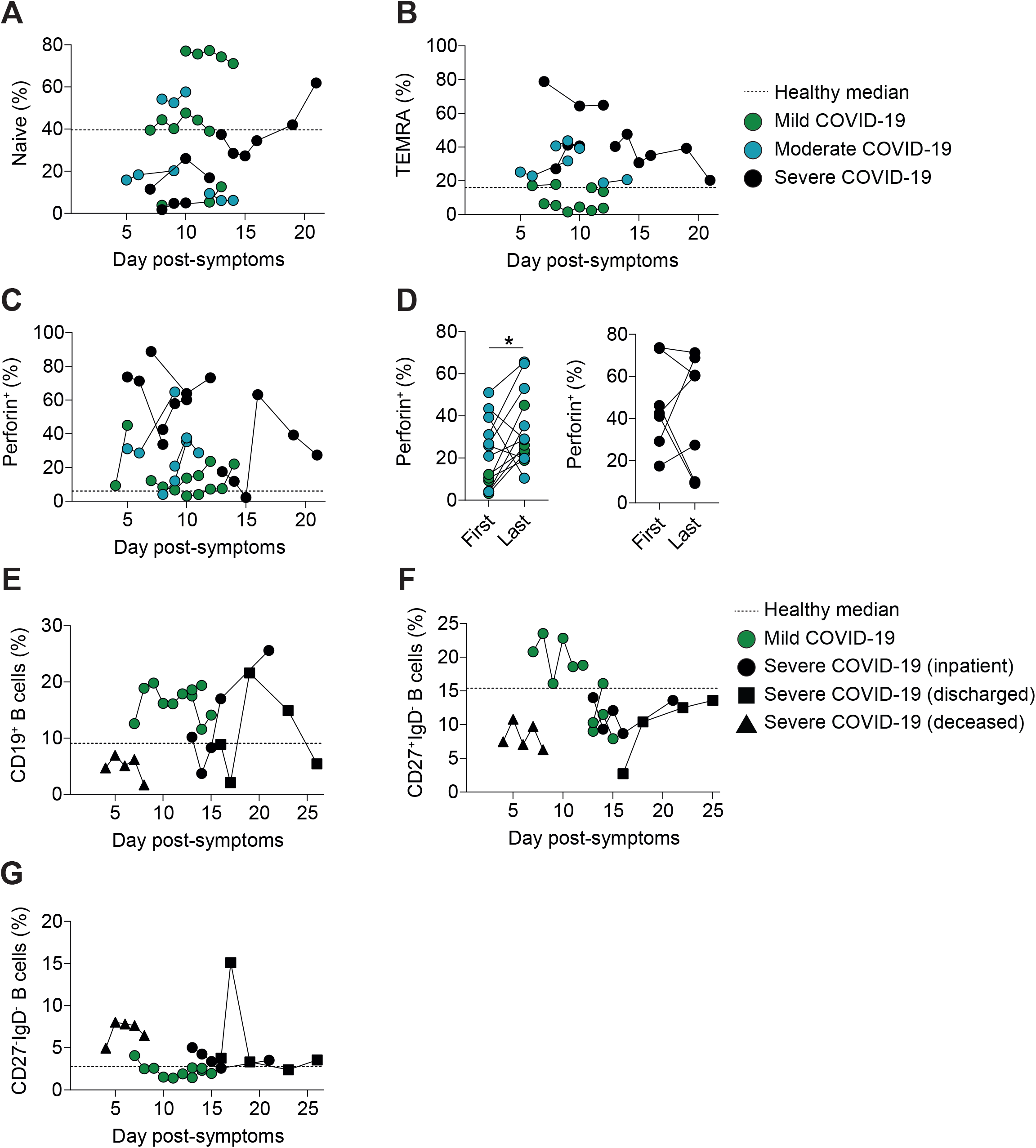

**Appendix Figure 8.**
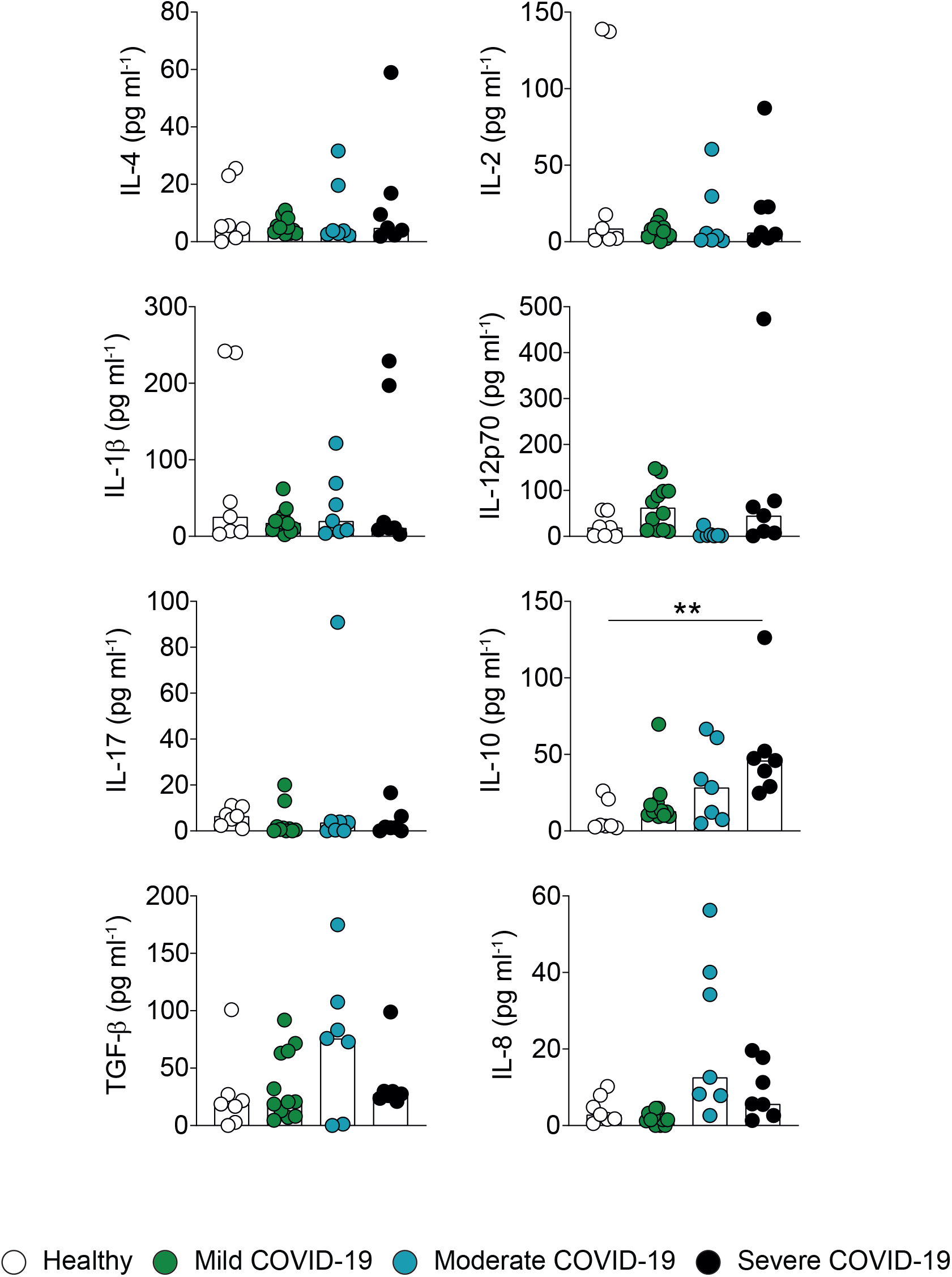

